# Differential Determinants of Past Behavior and Future Intention Regarding Voluntary Blood Donation: A Cross-Sectional Study of Knowledge, Attitudes, and Practices in Qingdao, China

**DOI:** 10.64898/2026.06.16.26355761

**Authors:** Feng Cheng, Li Zhang, Bohang Wang, Zining Dai

**Author notes:** Corresponding author Li Zhang (LZ).

## Abstract

**Background:** A persistent gap between motivation and action threatens voluntary blood supply. This study examined the public’s knowledge, attitudes, and practices (KAP) regarding blood donation, with a particular focus on identifying the different determinants of past blood donation behavior and future willingness to donate.

**Methods:** Convenience sampling was used to conduct a cross-sectional survey among 1,058 eligible people in Qingdao, China, between July and November 2025. Data were collected via a self-designed KAP questionnaire. To find independent characteristics linked to previous behavior and future intention, respectively, multivariable binary logistic regression was used.

**Results:** Overall, 37.0% of participants (n=391) had a lifetime donation history, while 39.2% (n=415) intended to donate in the next 12 months. Past behavior was positively associated with older age (36–45 years: OR=6.84; 95% CI: 3.21–14.58), higher education (OR=2.06; 95% CI: 1.33–3.17), and interpersonal interaction channels (OR=1.45; 95% CI: 1.01–2.09) but hindered by safety concerns (OR=0.23; 95% CI: 0.16–0.34). Conversely, future intention was positively correlated with male sex (OR=1.69; 95% CI: 1.24–2.29), prior donation history (OR=2.69; 95% CI: 1.87–3.86), having family members or friends in need of blood (OR=2.75; 95% CI: 1.96–3.85), and traditional media exposure (OR=3.33; 95% CI: 2.18–5.10). Higher education was adversely correlated with future intention (OR=0.55; 95% CI: 0.38–0.79).

**Conclusion:** There is a substantial disparity between donation motivation and action. The determinants of past behavior and future intention are asymmetric, suggesting that stage-specific interventions are required, using social mobilization for initiating first-time donations, while employing family reciprocity and authoritative communication to sustain long-term engagement.

**Author summary:** In this study, we investigated the knowledge, attitudes, and practices regarding voluntary blood donation among 1,058 eligible adults in Qingdao and found a notable “high motivation, low participation” paradox. More importantly, by contrasting logistic regression models, we identified that the determinants of past behavior and future intention are asymmetric—for instance, interpersonal interactions act as “initiating factors” for past behavior, while family reciprocity and traditional media exposure serve as “situational triggers” for future intention. These findings may provide actionable evidence to inform stage-specific, targeted recruitment and retention strategies, moving beyond traditional one-size-fits-all approaches. It directly addresses a critical global public health challenge, sustaining the voluntary blood supplyand provides practical insights for public health policy and intervention design.

## Introduction

Voluntary blood donation has become an essential public health concern globally, and is the foundation for a stable and safe blood supply for clinical therapies [1, 2]. Growing evidence suggests that demographic aging and the continuous advancement of medical technologies have progressively increased the clinical demand for blood products, making the recruitment and retention of eligible donors a high priority for healthcare systems worldwide [3, 4].

Previous studies have extensively investigated the Knowledge, Attitudes, and Practices (KAP) related to blood donation, identifying common barriers such as safety concerns, lack of awareness, and fear [5, 6]. Recent work has focused on the demographic and psychosocial determinants of donation behavior, demonstrating that factors such as altruistic motivation, social recognition, and interpersonal mobilization play significant roles in facilitating participation [7, 8].However, the current evidence remains limited in distinguishing the differential pathways leading to past donation behavior versus future donation intention [2]. Most existing literature treats behavior and intention as interchangeable endpoints or evaluates them in isolation. Few studies have examined whether the “initiating factors” that trigger first-time donations are congruent with the “maintaining factors” or “situational triggers” that sustain future willingness, particularly within rapidly urbanizing contexts such as Qingdao [9, 10]. Understanding this asymmetry is critical, as strategies effective for recruiting new donors may not necessarily retain existing ones [11].

Thus, the goal of the current study was to systematically investigate the KAP with reference to voluntary blood donation among Qingdao, China’s eligible population.Using cross-sectional survey data, we specifically sought to identify and contrast the differential determinants of previous donation behavior and future donation intention, thereby providing empirical evidence to inform targeted, stage-specific recruitment and retention strategies.

## Materials and methods

### Study Design

From July to November 2025, a cross-sectional study was carried out in the main urban region of Qingdao, China, to investigate the status of KAP with reference to voluntary blood donation. The design and reporting of this study complied with the requirements of the Strengthening of Reporting of Observational Studies in Epidemiology (STROBE) guidelines. The study protocol was approved by the Institutional Review Board of Qingdao Blood Center (Approval No. XZLL2025-002-01).

### Participants and Recruitment

Using a convenience sampling method, participants were recruited from multiple settings, including 5 enterprises and institutions, 3 residential communities, 3 universities, and selected blood donation houses in the main urban area of Qingdao, China. Eligible participants are individuals aged 18 to 60 who have signed an informed consent form and voluntarily agreed to enroll for this study.Individuals were excluded if they presented with visual, auditory, speech, reading, or cognitive impairments that would prevent them from completing the questionnaire independently, or if they had a history of psychiatric disorders.

### Measures and Data Collection

The data was acquired through a self-developed questionnaire designed based on a comprehensive systematic review and consultation with experts. The instrument, titled “Questionnaire on Blood Donation Behavior and Future Willingness to Donate Blood among the Population Aged 18-60 in Qingdao, China”, was structured based on the KAP theoretical framework and comprised six modules:

1. Demographic Characteristics: Sex, age group, educational level, occupation, and household registration (Hukou).
2. Knowledge and Policy Awareness: The assessment uses a 5-point Likert scale, with scores ranging from 1 (not at all familiar) to 5 (very familiar). The total scores for each dimension are calculated as the sum of the scores for each item.
3. Information Acquisition Channels: Including new media, traditional media, official channels, and interpersonal interactions.
4. Facilitators and Barriers: Items evaluating factors that promote or hinder blood donation (e.g., altruistic motivation, having family members or friends in need of blood, provision of personal emotional value, concerns about health and safety).
5. Previous Blood Donation Behavior: Defined as having donated blood at least once previously (coded as: 0 = never, 1 = ever).
6. Future Blood Donation Intention: Defined as the willingness to become a blood donor within the next 12 months (coded as: 0 = not willing, 1 = willing).

Following the initial draft, the questionnaire was reviewed and revised by three experts to ensure content validity. After the formal survey was completed, the internal consistency reliability for the blood donation knowledge and policy subscale was assessed. The results showed a Cronbach’s alpha coefficient of 0.939, indicating good reliability.

### Statistical Analysis

Categorical variables are described by frequency and percentage [n (%)], and comparisons between groups were performed using the chi-square test (χ²). Since the knowledge and policy scores did not follow a normal distribution, they are presented as medians and interquartile ranges (IQR).Multivariable binary logistic regression was conducted to determine the factors related to prior blood donation and the willingness to donate blood in the following 12 months. Variables with a *P* value below 0.10 in univariate analysis were included in the multivariable regression models and were further selected using the backward likelihood ratio method. The associations were expressed as odds ratios (ORs) and corresponding 95% confidence intervals (CIs). All analyses were carried out with SPSS software, version 26.0. Two-tailed tests were applied, and statistical significance was defined as a P value less than 0.05.

## Results

### Participants Characteristics

A total of 1,058 eligible questionnaires were collected. The demographic characteristics of the participants are shown in **Table Error! Reference source not found..**The sample was evenly distributed by sex (49.9% male vs. 50.1% female), and the majority held a bachelor’s degree or above (66.1%). Approximately one-third of participants (33.6%) were aged between 36 and 45 years. The lifetime cumulative prevalence of voluntary blood donation among respondents was 37.0% (n=391), while 39.2% (n=415) of respondents indicated an intention to donate blood sometime within the following 12 months. Univariate analysis revealed that age, educational level, and occupation were significantly associated with both past blood donation behavior and future donation intentions (all *P*< 0.001).

**Table 1.** Basic characteristics of participants and univariate analyses of blood donation behavior and intention (n=1058)

| Grouping | N(%) | Blood Donation Behavior |  |  |  | Intention to Donate Blood in the Next 12 Months |  |  |  |
| --- | --- | --- | --- | --- | --- | --- | --- | --- | --- |
| | | Never donated<br>n (%) | Ever donated<br>n (%) | $\chi^2$ | *p* value | No intention<br>n (%) | With<br>intention<br>n (%) | $\chi^2$ | *p* value |
| **Gender** |  |  |  |  |  |  |  |  |  |
| Male | 528 (49.9) | 328 (62.1) | 200 (37.9) | 0.385 | 0.535 | 282 (53.4) | 246 (46.6) | 23.989 | <0.001* |
| Female | 530 (50.1) | 339 (64.0) | 191 (36.0) |  |  | 361 (68.1) | 169 (31.9) |  |  |
| **Age group(years)** |  |  |  |  |  |  |  |  |  |
| 18-25 | 270 (25.5) | 217 (80.4) | 53 (19.6) | 66.538 | <0.001* | 202 (74.8) | 68 (25.2) | 53.317 | <0.001* |
| 26-35 | 286 (27.0) | 175 (61.2) | 111 (38.8) |  |  | 148 (51.7) | 138 (48.3) |  |  |
| 36-45 | 356 (33.6) | 175 (49.2) | 181 (50.8) |  |  | 230 (64.6) | 126 (35.4) |  |  |
| 46-60 | 146 (13.8) | 100 (68.5) | 46 (31.5) |  |  | 63 (43.2) | 83 (56.8) |  |  |
| **Educational level** |  |  |  |  |  |  |  |  |  |
| College or below | 359 (33.9) | 259 (72.1) | 100 (27.9) | 19.319 | <0.001* | 182 (50.7) | 177 (49.3) | 23.154 | <0.001* |
| Bachelor's or above | 699 (66.1) | 408 (58.4) | 291 (41.6) |  |  | 461 (66.0) | 238 (34.0) |  |  |
| **Occupation** |  |  |  |  |  |  |  |  |  |
| Professional and technicalworkers | 280 (26.5) | 109 (38.9) | 171 (61.1) | 106.078 | <0.001* | 170 (60.7) | 110 (39.3) | 49.132 | <0.001* |
| Students | 266 (25.1) | 212 (79.7) | 54 (20.3) |  |  | 207 (77.8) | 59 (22.2) |  |  |
| Others | 512 (48.4) | 346 (67.6) | 166 (32.4) |  |  | 266 (52.0) | 246 (48.0) |  |  |
| **Household registration** |  |  |  |  |  |  |  |  |  |
| Local or long-term residence (> 6Months) | 691 (65.3) | 437 (63.2) | 254 (36.8) | 0.034 | 0.855 | 408 (59.0) | 283 (41.0) | 2.501 | 0.114 |
| Non-local | 528 (34.7) | 230 (62.7) | 137 (37.3) |  |  | 235 (64.0) | 132 (36.0) |  |  |
| **Altruistic motivation** |  |  |  |  |  |  |  |  |  |
| selected | 858 (81.1) | 512 (59.7) | 346 (40.3) | 22.122 | <0.001* | 511 (79.5) | 347 (83.6) | 2.824 | 0.093* |
| Not selected |  | 155 (77.5) | 45 (22.5) |  |  | 132 (20.5) | 68 (16.4) |  |  |
| **Family members or friends in need of blood** |  |  |  |  |  |  |  |  |  |
| Selected | 372 (35.2) | 253 (68.0) | 119 (32.0) | 6.076 | 0.014* | 164 (25.5) | 208 (50.1) | 67.031 | <0.001* |
| Not selected |  | 414 (60.3) | 272 (39.7) |  |  | 479 (74.5) | 207 (49.9) |  |  |
| **Personal emotional value** |  |  |  |  |  |  |  |  |  |
| Selected | 327 (30.9) | 186 (56.9) | 141 (43.1) | 7.715 | 0.005* | 137 (21.3) | 190 (45.8) | 70.760 | <0.001* |
| Not selected |  | 481 (65.8) | 250 (34.2) |  |  | 506 (78.2) | 225 (54.2) |  |  |
| **Beneficial to own health and safety** |  |  |  |  |  |  |  |  |  |
| Selected | 265 (25.0) | 172 (64.9) | 93 (36.1) | 0.526 | 0.468 | 132 (20.5) | 133 (32.0) | 17.827 | <0.001* |
| Not selected |  | 495 (62.4) | 298 (37.6) |  |  | 511 (79.5) | 282 (68.0) |  |  |
| **Convenient time and location** |  |  |  |  |  |  |  |  |  |
| Selected | 180 (17.0) | 98 (54.4) | 82 (45.6) | 6.884 | 0.009* | 80 (12.4) | 100 (24.1) | 24.265 | <0.001* |
| Not selected |  | 569 (64.8) | 309 (35.2) |  |  | 563 (87.6) | 315 (75.9) |  |  |
| **Concerns about health and safety** |  |  |  |  |  |  |  |  |  |
| Selected | 763 (72.1) | 525 (68.8) | 238 (31.2) | 39.020 | <0.001* | 489 (76.0) | 274 (66.0) | 12.607 | <0.001* |
| Not selected |  | 142 (48.1) | 153 (51.9) |  |  | 154 (24.0) | 141 (34.0) |  |  |
| **Fear of blood, needles, or pain** |  |  |  |  |  |  |  |  |  |
| Selected | 300 (28.4) | 206 (68.7) | 94 (31.3) | 5.683 | 0.017* | 182 (28.3) | 118 (28.4) | 0.002 | 0.964 |
| Not selected |  | 461 (60.8) | 297 (39.2) |  |  | 461 (71.7) | 297 (71.6) |  |  |
| **Unaware of blood shortage and need for donation** |  |  |  |  |  |  |  |  |  |
| Selected | 230 (21.7) | 193 (83.9) | 37 (16.1) | 54.939 | <0.001* | 109 (17.0) | 121 (29.2) | 22.083 | <0.001* |
| Not selected |  | 474 (57.2) | 354 (42.8) |  |  | 534 (83.0) | 294 (70.8) |  |  |
| **Discouraged by family or friends** |  |  |  |  |  |  |  |  |  |
| Selected | 177 (16.7) | 123 (69.5) | 54 (30.5) | 3.793 | 0.051* | 111 (17.3) | 66 (15.9) | 0.334 | 0.563 |
| Not selected |  | 544 (61.7) | 337 (38.3) |  |  | 532 (82.7) | 349 (84.1) |  |  |
| **Concerns about blood stations profiting** |  |  |  |  |  |  |  |  |  |
| Selected | 123 (11.6) | 98 (79.7) | 25 (20.3) | 16.523 | <0.001* | 81 (12.6) | 42 (10.1) | 1.506 | 0.220 |
| Not selected |  | 569 (60.9) | 366 (39.1) |  |  | 562 (87.4) | 373 (89.9) |  |  |
| **Blood donation behavior** |  |  |  |  |  |  |  |  |  |
| Never donated | 667 (63.0) | - | - | - | - | 456 (68.4) | 211 (31.6) | 43.623 | <0.001* |
| Ever donated | 391 (34.0) | - | - |  |  | 187 (47.8) | 204 (52.2) |  |  |
| **Intention to donate blood in the next 12 months** |  |  |  |  |  |  |  |  |  |
| No intention | 643 (60.8) | - | - | - | - | - | - | - | - |
| With intention | 415 (39.2) | - | - |  |  | - | - |  |  |

### Knowledge and Policy Awareness

The median score for donation-related knowledge was 9.00 (IQR: 6.00–12.00), and the median score for policy awareness was 6.00 (IQR: 5.00–9.00) **(Table Error! Reference source not found.)**. Knowledge and policy scores were considerably higher among those who had donated blood in the past than among those who had not (*P*< 0.001). Similarly, participants with future donation intentions scored higher than those without such intentions (*P*< 0.001 for knowledge; *P* = 0.090 for policy).

**Table 2.**
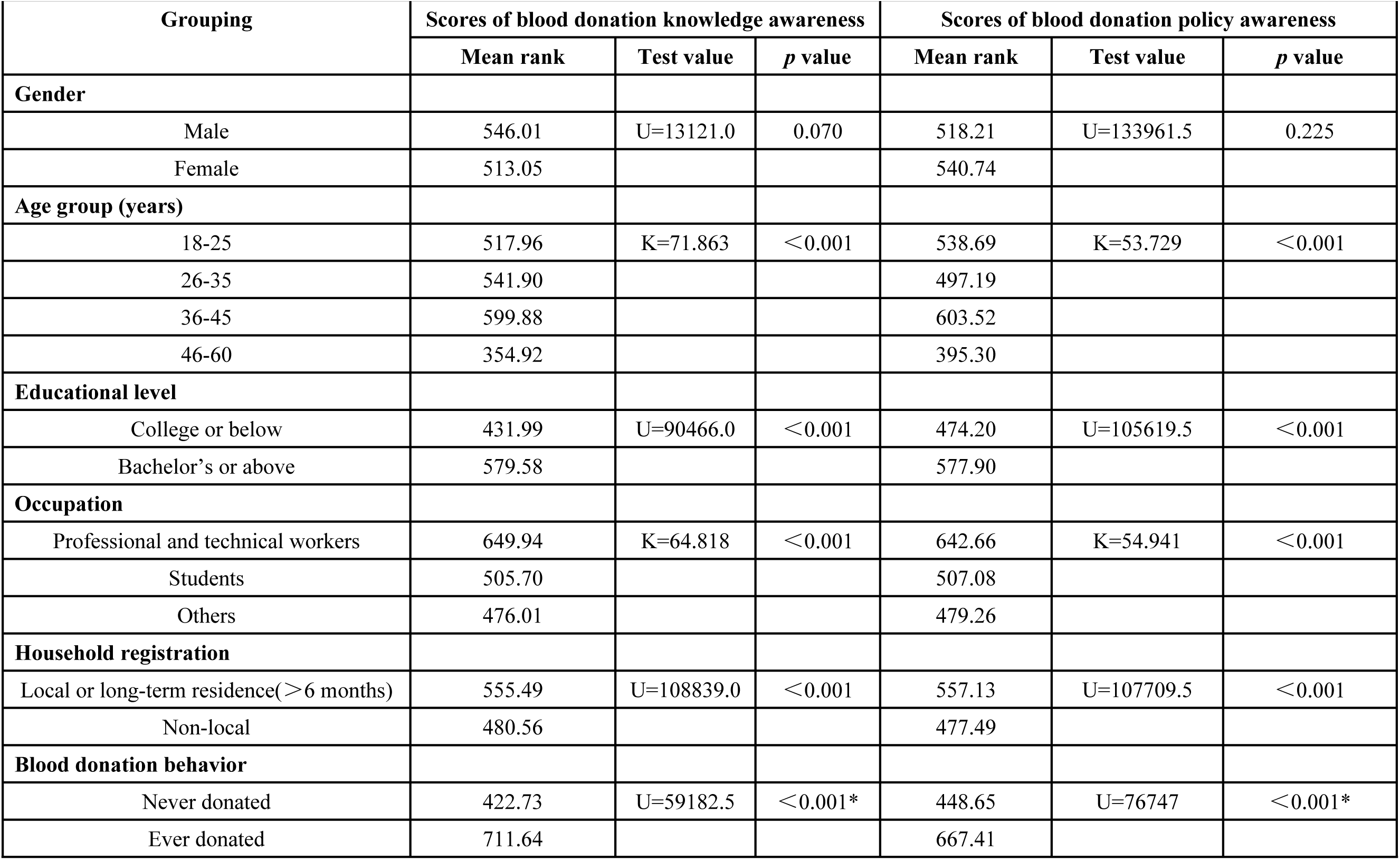
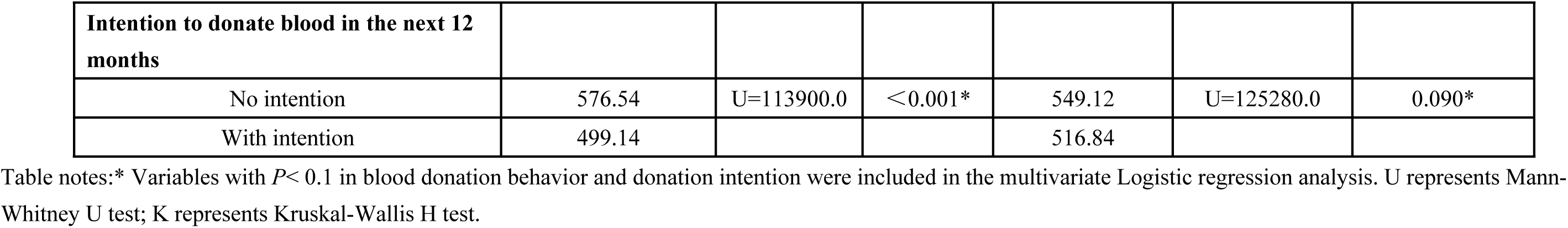
Comparison of blood donation knowledge and policy awareness scores among participants with different characteristics.

| Grouping | Scores of blood donation knowledge awareness |  |  | Scores of blood donation policy awareness |  |  |
| --- | --- | --- | --- | --- | --- | --- |
|  | Mean rank | Test value | *p* value | Mean rank | Test value | *p* value |
| **Gender** |  |  |  |  |  |  |
| Male | 546.01 | U=13121.0 | 0.070 | 518.21 | U=133961.5 | 0.225 |
| Female | 513.05 |  |  | 540.74 |  |  |
| **Age group (years)** |  |  |  |  |  |  |
| 18-25 | 517.96 | K=71.863 | <0.001 | 538.69 | K=53.729 | <0.001 |
| 26-35 | 541.90 |  |  | 497.19 |  |  |
| 36-45 | 599.88 |  |  | 603.52 |  |  |
| 46-60 | 354.92 |  |  | 395.30 |  |  |
| **Educational level** |  |  |  |  |  |  |
| College or below | 431.99 | U=90466.0 | <0.001 | 474.20 | U=105619.5 | <0.001 |
| Bachelor's or above | 579.58 |  |  | 577.90 |  |  |
| **Occupation** |  |  |  |  |  |  |
| Professional and technical workers | 649.94 | K=64.818 | <0.001 | 642.66 | K=54.941 | <0.001 |
| Students | 505.70 |  |  | 507.08 |  |  |
| Others | 476.01 |  |  | 479.26 |  |  |
| **Household registration** |  |  |  |  |  |  |
| Local or long-term residence(>6 months) | 555.49 | U=108839.0 | <0.001 | 557.13 | U=107709.5 | <0.001 |
| Non-local | 480.56 |  |  | 477.49 |  |  |
| **Blood donation behavior** |  |  |  |  |  |  |
| Never donated | 422.73 | U=59182.5 | <0.001* | 448.65 | U=76747 | <0.001* |
| Ever donated | 711.64 |  |  | 667.41 |  |  |
| **Intention to donate blood in the next 12 months** |  |  |  |  |  |  |
| No intention | 576.54 | U=113900.0 | <0.001* | 549.12 | U=125280.0 | 0.090* |
| With intention | 499.14 |  |  | 516.84 |  |  |

Table notes: \* Variables with $P < 0.1$ in blood donation behavior and donation intention were included in the multivariate Logistic regression analysis. U represents Mann- Whitney U test; K represents Kruskal-Wallis H test.

### Information Acquisition Channels

Regarding the channels through which participants acquired blood donation information **(Table Error! Reference source not found.)**, official channels were the most utilized (76.5%), followed by new media (61.1%) and interpersonal interactions (30.6%); traditional media were the least utilized (14.9%). Distinct demographic patterns emerged: younger participants and students were more likely to use new media (*P*< 0.001), whereas older and locally registered residents relied more on traditional media (*P*< 0.001). Individuals with previous donation experience were significantly more likely to obtain information via interpersonal interactions than those without experience (*P*< 0.001).

**Table 3.** Differences in participant characteristics by source of blood donation information.

| Grouping | New Media<br>Channel<br>n (%) | $\chi^2$ | *p* value | Traditional<br>Media<br>Channel<br>n (%) | $\chi^2$ | *p* value | Official<br>Channel<br>n (%) | $\chi^2$ | *p* value | Interpersona<br>l Interaction<br>Channel n<br>(%) | $\chi^2$ | *p* value |
| --- | --- | --- | --- | --- | --- | --- | --- | --- | --- | --- | --- | --- |
| **Total** | 646 (61.1) |  |  | 158 (14.9) |  |  | 809 (76.5) |  |  | 324 (30.6) |  |  |
| **Gender** |  |  |  |  |  |  |  |  |  |  |  |  |
| Male | 343 (65.0) | 6.755 | 0.009 | 130 (19.5) | 17.356 | <0.001 | 389 (73.7) | 4.562 | 0.033 | 150 (28.4) | 2.433 | 0.119 |
| Female | 303 (57.2) |  |  | 55 (10.4) |  |  | 420 (79.2) |  |  | 174 (32.8) |  |  |
| **Age group (years)** |  |  |  |  |  |  |  |  |  |  |  |  |
| 18-25 | 211 (78.1) | 59.306 | <0.001 | 41 (15.2) | 30.151 | <0.001 | 208 (77.0) | 0.430 | 0.934 | 93 (34.4) | 7.826 | 0.050 |
| 26-35 | 183 (64.0) |  |  | 32 (11.2) |  |  | 220 (76.9) |  |  | 76 (26.6) |  |  |
| 36-45 | 181 (50.8) |  |  | 42 (11.8) |  |  | 268 (75.3) |  |  | 119 (33.4) |  |  |
| 46-60 | 71 (48.6) |  |  | 43 (29.5) |  |  | 113 (77.4) |  |  | 36 (24.7) |  |  |
| **Educational Level** |  |  |  |  |  |  |  |  |  |  |  |  |
| College or below | 219 (61.0) | 0.001 | 0.979 | 63 (17.5) | 2.925 | 0.087 | 275 (76.6) | 0.006 | 0.940 | 117 (32.6) | 0.989 | 0.320 |
| Bachelor's or above | 427 (61.1) |  |  | 95 (13.6) |  |  | 534 (76.4) |  |  | 207 (29.6) |  |  |
| **Occupation** |  |  |  |  |  |  |  |  |  |  |  |  |
| Professional and<br>technical Workers | 151 (53.9) | 25.344 | <0.001 | 31 (11.1) | 6.326 | 0.042 | 235 (83.9) | 11.788 | 0.003 | 93 (33.2) | 1.628 | 0.443 |
| Students | 196 (73.7) |  |  | 37 (13.9) |  |  | 196 (73.7) |  |  | 75 (28.2) |  |  |
| Others | 299 (58.4) |  |  | 90 (17.6) |  |  | 378 (73.8) |  |  | 156 (30.5) |  |  |
| **Household**<br>**Registration** |  |  |  |  |  |  |  |  |  |  |  |  |
| Local or Long-Term Residence (>6 Months) | 398 (57.6) | 10.035 | 0.002 | 124 (17.9) | 14.218 | <0.001 | 538 (77.9) | 2.148 | 0.143 | 214 (31.0) | 0.112 | 0.738 |
| Non-Local | 248 (67.6) |  |  | 34 (9.3) |  |  | 271 (73.8) |  |  | 110 (30.0) |  |  |
| **Blood Donation Behavior** |  |  |  |  |  |  |  |  |  |  |  |  |
| Never Donated | 444 (66.6) | 23.029 | <0.001* | 121 (18.1) | 14.613 | <0.001* | 507 (76.0) | 0.206 | 0.650 | 178 (26.7) | 13.168 | <0.001* |
| Ever Donated | 202 (51.7) |  |  | 37 (9.5) |  |  | 302 (77.2) |  |  | 146 (37.3) |  |  |
| **Intention to Donate Blood in the Next 12 Months** |  |  |  |  |  |  |  |  |  |  |  |  |
| No Intention | 383 (59.6) | 1.539 | 0.215 | 60 (9.3) | 40.504 | <0.001* | 467 (72.6) | 13.409 | 0.000* | 191 (29.7) | 0.652 | 0.419 |
| With Intention | 263 (63.4) |  |  | 98 (23.6) |  |  | 342 (82.4) |  |  | 133 (32.0) |  |  |

Table note: \* Variables with $p < 0.1$ in blood donation behavior and donation intention were included in the multivariate Logistic regression analysis.

### Determinants of Past Blood Donation Behavior

Multivariable binary logistic regression identified several independent predictors of and barriers to previous blood donation behavior **(Table Error! Reference source not found.)**. Elderly age was confirmed to be a strong positive predictor, with the odds ratio increasing significantly across all age groups (26–35 years: OR = 3.585, 95% CI: 1.739–7.392; 36–45 years: OR = 6.839, 95% CI: 3.208–14.579; 46–60 years: OR = 11.061, 95% CI: 4.619–26.488; reference group: 18–25 years). Other significant positive predictors included holding a bachelor’s degree or higher (OR = 2.055, 95% CI: 1.332–3.169), working in a professional or technical role (OR = 2.516, 95% CI: 1.604–3.948), and higher knowledge scores (OR = 1.364, 95% CI: 1.285–1.447), acquiring information via interpersonal interactions (OR = 1.453, 95% CI: 1.011–2.089), and the desire to provide personal emotional value (OR = 1.523, 95% CI: 1.062–2.184). Conversely, concerns about health and safety (OR = 0.229, 95% CI: 0.155–0.338) and being unaware of blood supply shortages (OR = 0.216, 95% CI: 0.130–0.358) were significant barriers to past donation behavior.

**Table 4.** Logistic Regression Analysis of Blood Donation Behavior.

| | $\beta$ | SE | Wald | OR | 95% CI | *p* value |
| --- | --- | --- | --- | --- | --- | --- |
| **Age group (years)** |  |  |  |  |  |  |
| 26-35 | 1.277 | 0.369 | 11.957 | 3.585 | 1.739-7.392 | 0.001* |
| 36-45 | 1.923 | 0.386 | 24.788 | 6.839 | 3.208-14.579 | <0.001* |
| 46-60 | 2.403 | 0.446 | 29.098 | 11.061 | 4.619-26.488 | <0.001* |
| 18-25 |  |  |  | 1.000 |  |  |
| **Educational level** |  |  |  |  |  |  |
| Bachelor's or above | 0.720 | 0.221 | 10.614 | 2.055 | 1.332-3.169 | 0.001* |
| College or below |  |  |  | 1.000 |  |  |
| **Occupation** |  |  |  |  |  |  |
| Professional and technical workers | 0.923 | 0.230 | 16.127 | 2.516 | 1.604-3.948 | <0.001* |
| Students | 0.170 | 0.406 | 0.176 | 1.186 | 0.535-2.627 | 0.675 |
| Others |  |  |  | 1.000 |  |  |
| **Scores of blood donation knowledge awareness** | 0.310 | 0.030 | 104.631 | 1.364 | 1.285-1.447 | <0.001* |
| **Interpersonal interaction channel** |  |  |  |  |  |  |
| Selected | 0.374 | 0.185 | 4.068 | 1.453 | 1.011-2.089 | 0.044* |
| Not selected |  |  |  | 1.000 |  |  |
| **Personal emotional value** |  |  |  |  |  |  |
| Selected | 0.421 | 0.184 | 5.224 | 1.523 | 1.062-2.184 | 0.022* |
| Not Selected |  |  |  | 1.000 |  |  |
| **Concerns about health and safety** |  |  |  |  |  |  |
| Selected | -1.474 | 0.199 | 55.035 | 0.229 | 0.155-0.338 | <0.001* |
| Not selected |  |  |  | 1.000 |  |  |
| **Unaware of blood shortage and need for donation** |  |  |  |  |  |  |
| Selected | -1.533 | 0.257 | 35.485 | 0.216 | 0.130-0.358 | <0.001* |
| Not selected |  |  |  | 1.000 |  |  |
| **Constant** | -4.327 | 0.519 | 69.484 | 0.013 |  | <0.001* |

Table note: \*means $p < 0.05$ .

### Determinants of Future Blood Donation Intention

Male sex (OR = 1.686, 95% CI: 1.244–2.286) and having a previous donation history (OR = 2.686, 95% CI: 1.871–3.856) were prominent positive predictors **(Table Error! Reference source not found.)**. Similar to the behavior model, higher knowledge scores (OR = 1.087, 95% CI: 1.030–1.146) and the pursuit of personal emotional value (OR = 2.202, 95% CI: 1.593–3.045) positively predicted future intention. Information acquisition via traditional media (OR = 3.334, 95% CI: 2.179–5.101) and official channels (OR = 1.595, 95% CI: 1.125–2.262) were also strong facilitators. Among motivational factors, having family members or friends in need of blood transfusion was the strongest predictor (OR = 2.748, 95% CI: 1.964–3.846), followed by altruistic motivation (OR = 1.862, 95% CI: 1.242–2.791). Being unaware of blood supply shortages was associated with higher intention in this model (OR = 1.660, 95% CI: 1.129–2.441). Conversely, concerns about health and safety remained a significant barrier (OR = 0.674, 95% CI: 0.482–0.943). Holding a bachelor’s degree or above was inversely associated with future intention (OR = 0.548, 95% CI: 0.382–0.788), the opposite direction to its effect on past behavior.

**Table 5.** Logistic Regression Analysis of Intention to Donate Blood in the Next 12 Months.

| | $\beta$ | SE | Wald | OR | 95% CI | *p* value |
| --- | --- | --- | --- | --- | --- | --- |
| **Gender** |  |  |  |  |  |  |
| Male | 0.523 | 0.155 | 11.318 | 1.686 | 1.244-2.286 | 0.001* |
| Female |  |  |  | 1.000 |  |  |
| **Age group (years)** |  |  |  |  |  |  |
| 26-35 | 0.917 | 0.221 | 17.145 | 2.502 | 1.621-3.862 | <0.001* |
| 36-45 | 0.176 | 0.226 | 0.609 | 1.193 | 0.766-1.856 | 0.435 |
| 45-60 | 1.008 | 0.285 | 12.523 | 2.741 | 1.568-4.791 | <0.001* |
| 18-25 |  |  |  | 1.000 |  |  |
| **Educational level** |  |  |  |  |  |  |
| Bachelor's or above | -0.601 | 0.185 | 10.558 | 0.548 | 0.382-0.788 | 0.001* |
| College or below |  |  |  | 1.000 |  |  |
| **Blood donation behavior** |  |  |  |  |  |  |
| Ever Donated | 0.988 | 0.185 | 28.653 | 2.686 | 1.871-3.856 | <0.001* |
| Never Donated |  |  |  | 1.000 |  |  |
| **Scores of blood donation knowledge awareness** | 0.083 | 0.027 | 9.335 | 1.087 | 1.030-1.146 | 0.002* |
| **Traditional media channel** |  |  |  |  |  |  |
| Selected | 1.204 | 0.217 | 30.783 | 3.334 | 2.179-5.101 | <0.001* |
| Not selected |  |  |  | 1.000 |  |  |
| **Official channel** |  |  |  |  |  |  |
| Selected | 0.467 | 0.178 | 6.877 | 1.595 | 1.125-2.262 | 0.009* |
| Not selected |  |  |  | 1.000 |  |  |
| **Altruistic motivation** |  |  |  |  |  |  |
| Selected | 0.622 | 0.207 | 9.053 | 1.862 | 1.242-2.791 | <0.001* |
| Not selected |  |  |  | 1.000 |  |  |
| **Personal emotional value** |  |  |  |  |  |  |
| Selected | 0.790 | 0.165 | 22.820 | 2.202 | 1.593-3.045 | <0.001* |
| Not selected |  |  |  | 1.000 |  |  |
| **Family members or friends in need of blood** |  |  |  |  |  |  |
| Selected | 1.011 | 0.171 | 34.766 | 2.748 | 1.964-3.846 | <0.001* |
| Not selected |  |  |  | 1.000 |  |  |
| **Concerns about health and safety** |  |  |  |  |  |  |
| Selected | -0.394 | 0.171 | 5.296 | 0.674 | 0.482-0.943 | 0.021* |
| Not selected |  |  |  | 1.000 |  |  |
| **Unaware of blood shortage and need for donation** |  |  |  |  |  |  |
| Selected | 0.507 | 0.197 | 6.630 | 1.660 | 1.129-2.441 | 0.010* |
| Not selected |  |  |  | 1.000 |  |  |
| **Constant** | -3.396 | 0.397 | 73.327 | 0.034 |  | <0.001* |

Table note:\*means $p < 0.05$ .

## Discussion

The knowledge, attitudes, and practices of eligible individuals in Qingdao, China, about voluntary blood donation were systematically investigated in this study. We also identified the differential determinants of previous donation behavior and future donation intention. Our findings reveal a notable “high motivation, low participation” paradox: while over 80% of respondents reported altruistic motivations, the actual lifetime prevalence of donation was only 37.0%, and the willingness to donate in the following 12 months was 39.2%. This disparity suggests a disconnect in the transition from positive attitudes to concrete actions, consistent with prior research showing that intention frequently fails to translate into donation behavior [12, 13]. By contrasting the two logistic regression models, our study extends the current literature by distinguishing “initiating factors” (e.g., interpersonal interactions, professional background) that drive first-time behavior from “maintaining or situational triggers” (e.g., family need, traditional media exposure) that sustain future intention.

The differential effects of demographic characteristics indicate the heterogeneity of the donation pathway. Consistent with some recent multi-center studies, older age emerged as a stable independent predictor of both behavior and intention [14]. This contrasts with the traditional assumption that younger adults are the primary donor base [15]. An alternative explanation may be the concept of health privilege, wherein older adults, enjoying better health and having more flexible time, feel a stronger imperative to help others [16, 17]. Education exhibited a paradoxical dual effect, such that higher education facilitated past behavior but deterred future intention [12, 18]. While advanced education may enhance knowledge and confidence in the medical system initially, it may also increase opportunity costs and increase risk aversion regarding personal health and safety, thereby suppressing the willingness to donate again [2]. Similarly, gender significantly influenced intention but not past behavior, suggesting that while males may express a stronger helper identity, actual donation behavior may be more contingent on situational opportunities than on sex itself [19, 20].

Our findings highlight the distinct roles of information channels in shaping behavior versus intention. Exposure through interpersonal interactions was positively associated with past behavior, supporting the notion that social network mobilization is critical for the consolidation of donation behavior [21]. Conversely, traditional media and official channels was primarily associated with future intention. The authority and scientific credibility conveyed by these sources may help reduce uncertainty and alleviate safety concerns, thereby reducing hesitancy and increasing willingness [22, 23]. However, whether this intention translates into actual behavior warrants longitudinal verification [22].

Psychological motivations and barriers further define the scope of the KAP model. Providing personal emotional value consistently promoted both behavior and intention, indicating that donation is not merely an act of pure altruism, but a reciprocal social action embedded with “emotional return” and “social recognition”[24, 25]. This provides supportive evidence for policies such as China’s honor incentives, which function as symbols of social acknowledgment rather than economic compensation [26]. Furthermore, “family or friends needing blood” served as a strong situational trigger for intention, shifting the motivation from generalized helping to fulfilling relational responsibilities [26]. Conversely, health and safety concerns constituted a persistent barrier across both models, reaffirming them as the core barrier throughout the donation continuum [27, 28]. Providing personal emotional value was consistently positively associated with both behavior and intention. This may reflect a recruitment bias, whereby respondents surveyed at donation sites who were initially unaware might have been persuaded by the on-site atmosphere, generating an immediate but perhaps transient willingness.

This study has several limitations. First, its cross-sectional design does not allow causal inference, and the associations observed should therefore be interpreted as preliminary rather than definitive. Second, as participants were recruited using convenience sampling primarily from donation sites, the findings may not be fully generalizable to the broader urban population. Such a recruitment strategy may also have introduced volunteer bias, potentially leading to an overestimation of donation intention. Third, the assessment of intention to donate in the future was based on self-report, making it vulnerable to social desirability bias and limiting its ability to predict actual future behavior. Finally, residual confounding cannot be excluded despite adjusting for major demographic variables.

## Conclusion

In summary, this study discovered a substantial discrepancy between voluntary blood donation behavior and motivation. The determinants of past behavior and future intention are asymmetric, suggesting that targeted interventions must differentiate between strategies aimed at initiating first-time donations and those designed to sustain long-term engagement. Moving beyond traditional health education, future mobilization strategies should incorporate multidimensional approaches using social identity, family reciprocity, and emotional motivation, while concurrently addressing persistent safety concerns through authoritative, transparent communication.

## Data Availability

The minimal data set and accompanying code generated in this study are available at Zenodo via https://doi.org/10.5281/zenodo.20659632

## Acknowledgement

We would like to express our sincere gratitude to Dr. Zhan Su from The Affiliated Hospital of Qingdao University and Dr. Wei Yin from Guangzhou Medical University for their meticulous guidance and valuable advice during data analysis and manuscript preparation. We also appreciate the active participation and cooperation of all voluntary blood donors involved in this study.

